# Efficacy of SARS-CoV-2 vaccine in thoracic cancer patients: a prospective study supporting a third dose in patients with minimal serologic response after two vaccine doses

**DOI:** 10.1101/2021.08.12.21261806

**Authors:** Valérie Gounant, Valentine Marie Ferré, Ghassen Soussi, Charlotte Charpentier, Héloïse Flament, Nadhira Fidouh, Gilles Collin, Céline Namour, Sandra Assoun, Alexandra Bizot, Zohra Brouk, Eric Vicaut, Luis Teixeira, Diane Descamps, Gérard Zalcman

**Author notes:** To whom correspondence must be addressed: Prof. **G. Zalcman** Service d’ Oncologie Thoracique Hôpital Bichat-Claude Bernard 46 rue Henri Huchard FR-75018 Paris. both authors equally contributed (Valérie Gounant, Valentine Marie Ferré). **Authors contribution:***conceptualization*: GZ, VG, DD, LT; supervision: GZ, VG; *data curation*: GZ, CM, ZB, GS, VG, VMF; *Formal analysis:* GZ, GS, VG, EV, VMF; *Funding acquisition:* GZ, LT, HF, DD, EV; *Investigation:* GZ, VG, GS, SA, AB, CM, ZB; *Biological analyses:* VMF, CC, HF, NF, GC, DD; *Statistical analyses:* GS, EV; *original draft:* GZ; review & *editing:* GZ, VG, GS, VMF, HF DD, EV, EV; *final MS approbation:* all authors. Conflict of interest statement: Valentine Marie Ferré, Ghassen Soussi, Charlotte Charpentier, Héloïse Flament, Nadhira Fidouh, Gilles Collin, Céline Namour, Sandra Assoun, Alexandra Bizot, Zohra Brouk, Eric Vicaut, Luis Teixeira have nothing to disclose. **Gérard Zalcman** received research grants from Roche-France, BMS, Takeda outside of the submitted work, perceived fees from BMS, Astra-Zeneca, Pfizer, Borhinger-Ingelheim outside the submitted work and reimbursement for international meetings assistance from Abbvie, MSD, Astra-Zeneca, BMS, Roche-France; **Valérie Gounant** reports personal fees from MSD, Chugai, Novartis, and Boehringer; personal fees and non-financial support from Astra Zeneca, BMS, Takeda, and Pfizer; grants, personal fees, and non-financial support from Roche, all outside the submitted work; **Diane Descamps** reports fees from Viiv-healthcare, Gilead-Sciences, Janssen-Cilag outside the submitted work.

## Abstract

**Hypothesis:** Coronavirus disease 2019 (COVID-19) resulted in a 30% mortality rate in thoracic cancer patients. Given that cancer patients were excluded from serum anti-severe acute respiratory syndrome coronavirus 2 (SARS-CoV2) vaccine registration trials, it is still unknown whether they would develop a protective anti-spike antibody response following vaccination. This prospective vaccine monitoring study primarily aimed to assess humoral responses to SARS-CoV2 vaccine in thoracic cancer patients.

**Methods:** SARS-CoV2-spike antibodies were measured using Abbot ARCHITECT SARS-CoV-2 IgG immunoassay, prior to first injection of BNT162b2 mRNA vaccine, as well as at Week 4, and two-to-sixteen weeks after second vaccine dose. The factors associated with antibody response were analyzed.

**Results:** Overall, 306 patients, with a median age of 67.0 years (IQR=58-74), were vaccinated. Of these, 283 patients received two vaccine doses at 28-day intervals. After 4.7-month median follow-up, seven patients (2.3%) contracted proven symptomatic SARS-CoV-2 infection, with rapid favorable evolution. Of 269 serological results available beyond Day 14 post-second vaccine dose, 17 (6.3%) were still negative (<50 AU/mL) (arbitrary units/mL), while 34 (11**%)** were <300 AU/mL (12.5^th^ percentile). In multivariate analysis, only age and chronic corticosteroid treatment were significantly associated with a lack of immunization. Thirty patients received a third vaccine dose, with only three patients showing persistent negative serology thereafter, whereas the others demonstrated clear seroconversion.

**Conclusion:** SARS-CoV2 vaccines were shown to be efficient in thoracic cancer patients, most of them being immunized after two doses. A third shot given to 1% of patients with persistent low antibody titers resulted in a 88% immunization rate.

## Introduction

Coronavirus disease 2019 (COVID-19) is associated with a dramatic 30% mortality rate in thoracic cancer patients^1,2,3^. The Chinese series reported mortality rates of 29-39%,^4–6^ compared with 0.7-8.0% case fatality rates in their general population^7–11^. Lung cancer patients should therefore be given priority for severe acute respiratory syndrome coronavirus 2 (SARS-CoV-2) vaccination. Nonetheless, active cancer condition and immunosuppressive therapy constituted non-inclusion criteria for SARS-CoV-2 vaccine registration trials, and scarcely anything is known about the vaccine effectiveness in cancer populations. Moreover, the antibody response after influenza vaccination was previously shown to be lower in cancer patients versus healthy controls, especially concerning people aged ≥65 years^12^. Notably, two doses of influenza vaccine were required in cancer patients to attain the same serum protection rate than in healthy controls, with on-going chemotherapy or corticosteroids resulting in lower protection^13^. Similarly, a meta-analysis on influenza vaccine effectiveness in cancer patients exhibited significantly reduced seroconversion (≥four-fold rise) in comparison with vaccinated immunocompetent controls (0.31; 95%CI:0.22-0.43)^14^. Vaccination timing remains unclear too. Upon chemotherapy, the mid-point between two cycles was empirically selected for the vaccine shot. Moreover, as immunotherapy has become an essential component of lung cancer treatment, only little is known concerning undesirable effects, although no short-term reactogenicity following influenza vaccination occurred in patients under immune checkpoints inhibitors (ICIs)^15^. Both mRNA vaccine registration trials reported 94% effectiveness against severe COVID19 in healthy volunteers, ≥14 days post-second booster (Day 21 for Pfizer BNT162b2^16^ or Day 28 for Moderna mRNA-1273^17, 18^). Concerning adenovirus-based vaccines (one shot of Janssen Ad26.COV2.S^19^ or two doses of Astra-Zeneca ChAdOx1nCov-19 vaccines^20, 21^), slightly lower rates were reported. Similar protection rates against severe COVID19 were confirmed in real-life by population-based Israeli and Scottish studies for Pfizer BNT162b2 or ChAdOx1nCov-19^22, 23^. Yet, these data cannot be extrapolated to cancer patients undergoing anti-cancer treatment. Duration of anti-SARS-CoV-2-spike (anti-S) detection was at least 8 months in healthy volunteers^24^. It is unclear whether such duration is applicable to cancer patients receiving immunosuppressing drugs. Early January 2021, vaccination was made available in France. To increase first vaccine dose availability, French Health authorities recommended a 28-day interval for both mRNA vaccines, with a 42-day interval for healthy people, this delayed second dosing being debatable^25, 26^. Due to uncertainties concerning vaccination of cancer patients, the observational COVIDVAC-OH study (clinical.trials.gov NCT04776005) was launched, sponsored by Paris University Hospitals. This study sought to investigate SARS-CoV-2 vaccination’s (mainly mRNA-based vaccines) effectiveness in over 1100 consecutive patients with solid cancers or hematological malignancies, at North-Paris University Cancer center. This report concerns 306 thoracic cancer patients.

## METHODS

### Trial design, objectives, and participants

We conducted a prospective study involving thoracic cancer patients followed-up in Bichat Hospital, from January 26 to July 28, 2021. Patients diagnosed with thoracic cancer and deemed eligible (no known COVID-19 infection within <last three months; life expectancy >3 months; lack of known allergy to previous vaccines) were identified from medical records. They were contacted and offered to be vaccinated. If they accepted and in the absence of contra-indications, they were attended vaccination sessions in the outpatient clinic, according to priority sequencing, as follows: first elderly patients aged ≥75 years and those receiving chemotherapy; patients receiving immune checkpoint inhibitors (ICI); patients with pneumonectomy or chronic radiation pneumonitis; patients on oral tyrosine kinase targeted therapy (TKIs); patients without systemic therapy. They were given a written information leaflet on COVID19 mRNA BNT162b2 vaccine, and on serological and hematological blood tests to be performed at first dose (Day 0), at second dose (Day 28), and at least 2 weeks post-booster dose (Day 42). All patients could oppose blood samplings and still undergo vaccine injections. Recommendations to keep facial masks and social distancing were given. All patients were registered into National Health Insurance computed COVID-19 vaccine database, with national identification number, complete identity, main underlying conditions, and vaccine batch number.

Following blood sampling and vaccine injection, patients were followed-up for 15 minutes under medical surveillance^27^. This study was approved by the Comité d’Evaluation de l’Ethique des projets de Recherche Biomedicale (CEERB, IRB00006477) of University Hospitals Paris-North Val-de-Seine, Paris 7 University, Assistance Publique-Hôpitaux de Paris (AP-HP) upon the approval number N°CER-2021-72. In the case of COVID-19-suggestive symptoms, patients were instructed to promptly inform the medical team and perform nasopharyngeal SARS-CoV-2 RT-PCR swab testing. At second vaccination visit, they were questioned about undesirable events post-first vaccination and symptoms evoking COVID-19. Some patients (n=16) were vaccinated by their general practitioner or in a government-certified vaccination center. No blood sampling was available for these patients at Days 0 and 28. If they agreed, they underwent blood sampling in the post-booster period and were included in our study.

To set up the technical conditions for SARS-CoV-2 anti-nucleocapsid (N) index and anti-Spike antibody determination, 18 control subjects from Hôpital Bichat staff, without previous COVID-19 symptoms or PCR-proved SARS-CoV-2 infection, provided their written consent for blood sampling. This occurred on Day 28 post-first vaccine, while 13 subjects underwent additional blood sampling at least 2 weeks post-second vaccine.

### Study endpoints

The primary endpoint was to assess humoral immunity against SARS-CoV-2-spike in thoracic cancer patients following COVID-19 mRNA BNT162b2 vaccine injection and booster dose. Some patients vaccinated outside our center received Moderna mRNA-1273 (n=1) or Astra-Zeneca ChAdOx1 nCov-19 (n=3) vaccines, being included herein.

Secondary endpoints were vaccination safety, clinical efficacy based on RT-PCR-documented COVID-19 infection during the study, and hospitalization or death from COVID-19. Phone safety consultations were scheduled every 3 weeks. Cell immunity to SARS-CoV-2-spike protein was evaluated using T-cell enzyme linked immunospot (ELISPOT), with lymphocyte subset counts, scheduled at Day 28 and from Day 42 post-first injection in 122 arbitrarily-designated patients.

### Laboratory analyses

SARS-CoV-2 anti-N and anti-S antibody titers were determined using Abbott Architect SARS-CoV-2 IgG and IgG Quant II (Abbott, Maidenhead, UK) and expressed as index (cut-off: 0.49) and arbitrary units (cut-off: 50 AU/mL), respectively. Pseudo-neutralization assay was performed using iFlash-2019-nCoV Nab assay (YHLO, Shenzhen, China), which assesses antibody neutralizing capacity by competition with angiotensin-converting enzyme 2 (ACE2)-receptor for binding to anti-spike RBD (cut-off: 10AU/mL). This assay correlated with SARS-CoV-2 *in vitro* cell micro-neutralization (paper submitted for publication).

#### ELISspot assay methods are in online supplement

##### Statistical analysis

All samples were de-identified and assigned an ID number, with the sampling date. Sample processing and data analyses were performed, with all study personal blinded to information concerning patients and samples. De-identified data were exported from Microsoft Excel Version 2013 for Windows (Microsoft Corporation, 2013) to IBM® SPSS® Statistics for Windows, Version 25.0 (IBM Corp., Armonk, N.Y., USA) for statistical analysis. Between-group comparisons were performed using Pearson’s chi-squared or Fisher’s exact tests for discrete variables, and Student’s-T or Mann-Whitney U tests for continuous variables. Odds ratios (OR) and respective 95% confidence intervals (95%CI) were calculated using binary logistic regression. Hypothesis testing was two-tailed, with p values<0.05 considered statistically significant. All assumptions required for logistic regression modeling were verified. Multivariable analysis was conducted using binary logistic regression using the Enter method, including variables exhibiting a significance threshold p<0.20 yielded by the univariable analysis..

##### Funding source

The academic authors retained editorial control. The *Assistance Publique-Hôpitaux de Paris* funded the study, without participating to study design, data collection, data analysis, data interpretation, or report writing.

## RESULTS

### Participant characteristics

From January 20 to June 1, 2021, overall 325 thoracic cancer patients followed-up in thoracic oncology/surgery departments were proposed anti-SARS-CoV-2 vaccination with Pfizer BNT162b2 mRNA vaccine. Initially, only 36 **(11%)** declined the proposal. Of these, **17** eventually accepted to be vaccinated; nine of whom were vaccinated outside of our center but participated to serological testing. Overall, 306 patients received their 28-day-spaced doses or underwent blood samplings between January 26 and May 17, 2021. Patient disposition is illustrated in **suppl. Figure 1.** Clinical follow-up was extended until July 28, 2021.

**Figure 1:**
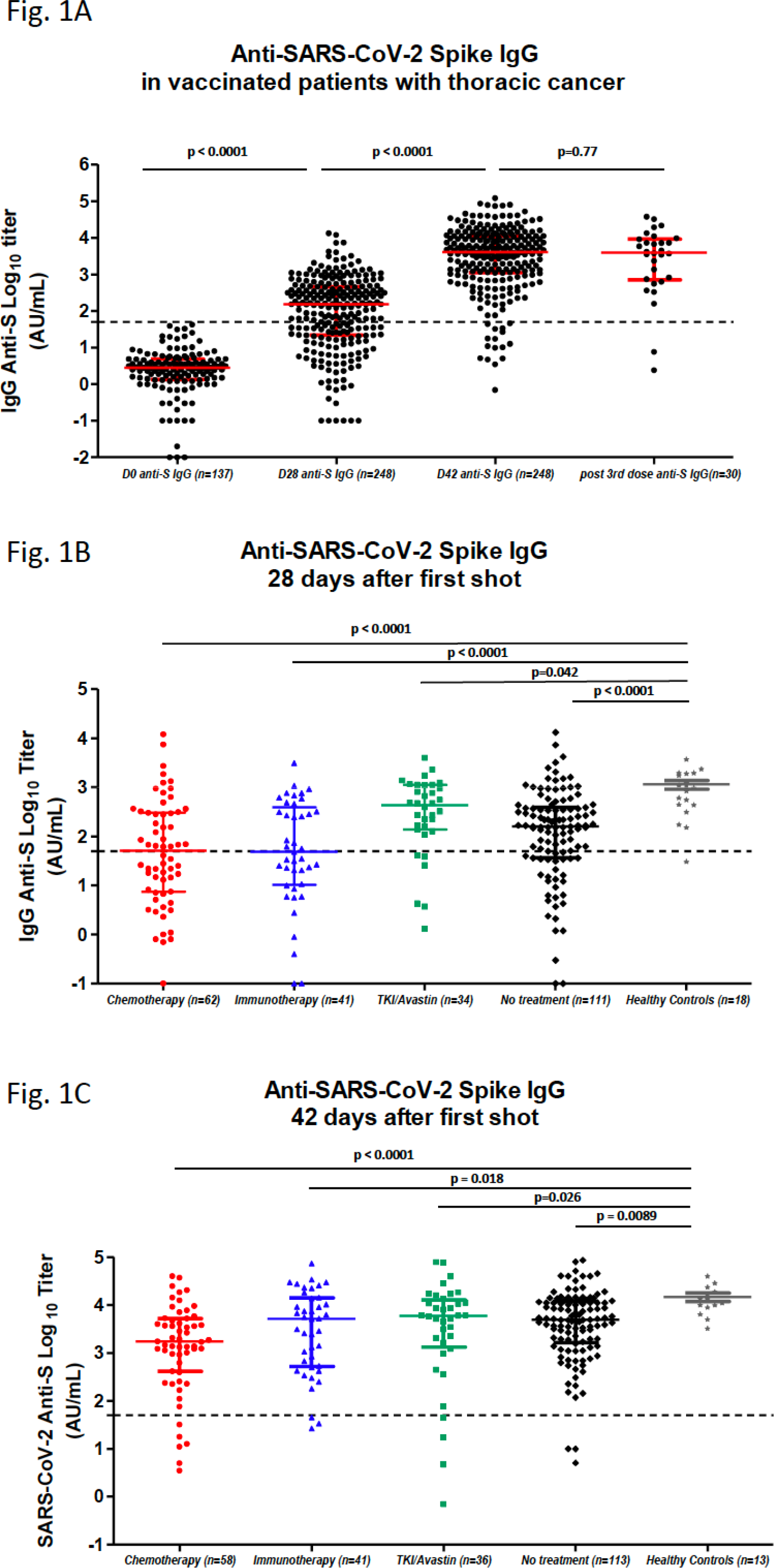
Serological response to COVID-19 vaccine BNT162b2 in COVID-free Patients **(A)** Anti-SARS-CoV-2-spike (anti-S) IgG antibody titers at Day 0 in 137 patients without previous history of COVID-19; at Day 28 after one vaccine dose injection in 248 patients without previous history of COVID-19; beyond Day 42 in 248 patients without previous history of COVID-19; beyond Day 21 after third vaccine dose in 30 patients with available results. Large horizontal bars represent the median value, with short bars showing the first (lower) and third (upper) quartiles values. The Mann-Whiney test was applied for statistical comparison. **(B)** Anti-S IgG antibody titers at Day 28 after the first vaccine dose, according to the systemic treatment received within the previous three months: chemotherapy, including chemo-immunotherapy (n=62), immunotherapy alone (n=41), oral tyrosine kinase inhibitor (TKI) or bevacizumab single-agent therapy (n=34), or without systemic treatment (n=111). Anti-S IgG antibody titers at Day 28 in 18 healthy controls are shown. Large horizontal bars represent the median value, with short bars showing the first (lower) and third (upper) quartiles values. The Mann-Whiney test was applied for statistical comparison. **(C)** Anti-S IgG antibody titers at Day 42 or beyond after the first vaccine dose, according to the systemic treatment received within the previous three months: chemotherapy, including chemo-immunotherapy (n=58), immunotherapy alone (n=41), oral TKI or bevacizumab single-agent (n=36), or without systemic treatment (n=113). Anti-SARS-CoV-2-spike IgG antibody titers at Day 42 in 13 healthy controls were available. Large horizontal bars represent the median value, with short bars showing the first (lower) and third (upper) quartiles values. The Mann-Whiney test was applied for statistical comparison.

Patient clinical and demographic characteristics are provided in **Table 1**. Overall, 181 patients (59.2%) were male and 285 (93.1%) exhibited lung cancer with 260 (84.9%) non-small cell lung carcinomas (NSCLCs) and 22 (7.2%) small-cell lung carcinomas (SCLCs), while 13 (4.4%) suffered from pleural malignant mesothelioma. Median age was 67 years IQR(58-74), with 41.2 % older than 70. Most patients (57.2%) displayed late-stage disease, with 117 (38%) cancer-diagnosed within <last 12 months. Last treatment received within three months prior-first vaccine was chemotherapy (n=74; 24.2 %), given alone (51; 16.7%), with concurrent thoracic radiotherapy (n=2), or combined with ICI (21, 6.9%), while 49 patients (16%) received ICI alone, and 13.7% were treated with daily TKIs or maintenance bevacizumab. The last 141 patients (30.7%) had not received systemic treatment within last three months. Overall, **37** (12.1%) patients displayed chronic radiation pneumonitis following radio-chemotherapy of Stage III lung cancer. A thoracic surgery history was recorded in 89 (29%) patients, six of whom (1.95%) underwent pneumonectomy and 79 (25.8%) lobectomy or sublobar resection. There were 20 (6.5%) patients under oral corticosteroids for at least 3 weeks, for immune-mediated ICI toxicity, pain, brain metastasis, or severe chronic obstructive pulmonary disease. Overall, 59.5% patients were in complete or partial response at vaccination time, while 29.1% were only recently cancer-diagnosed or displayed progressive disease.

**Table 1:**
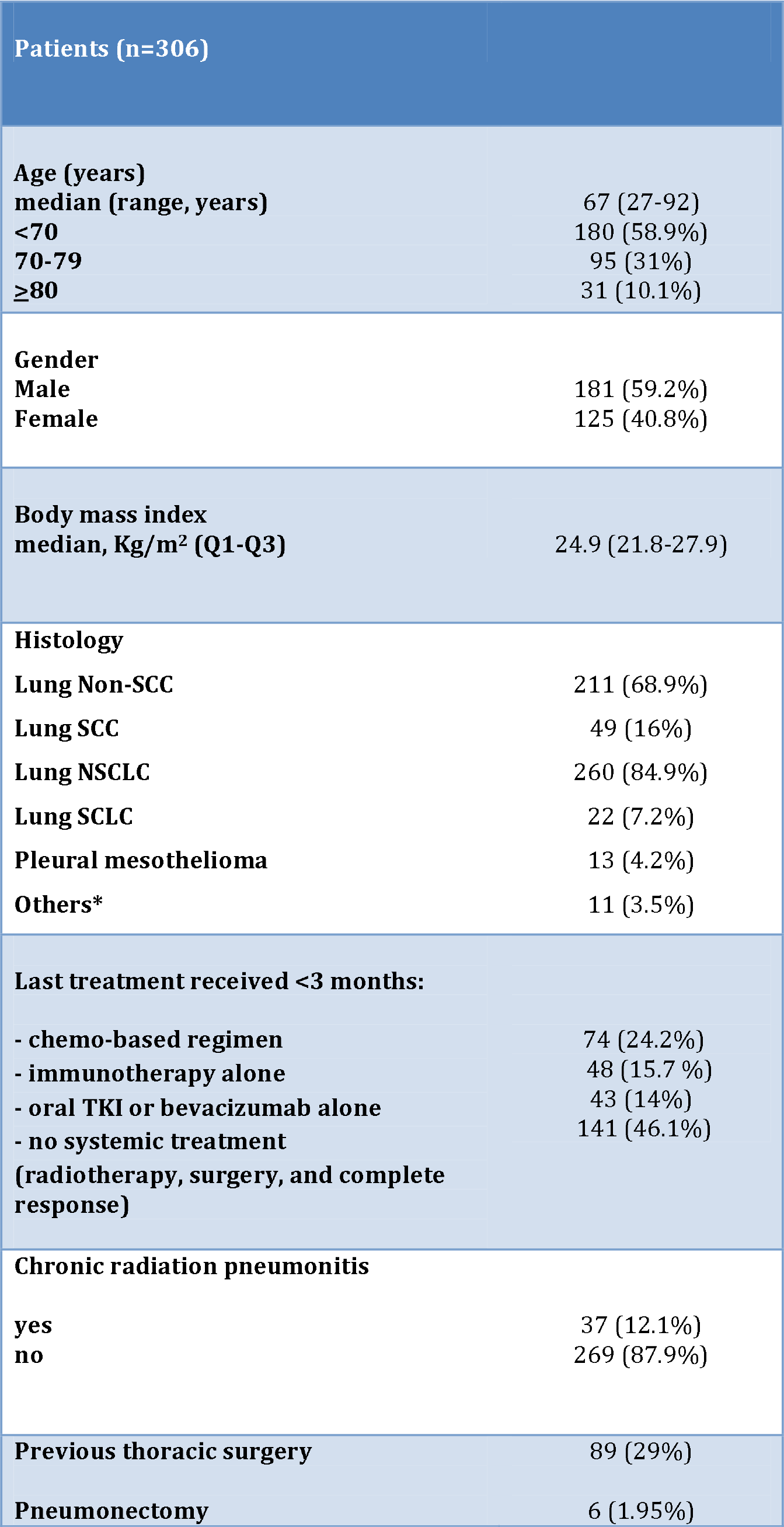
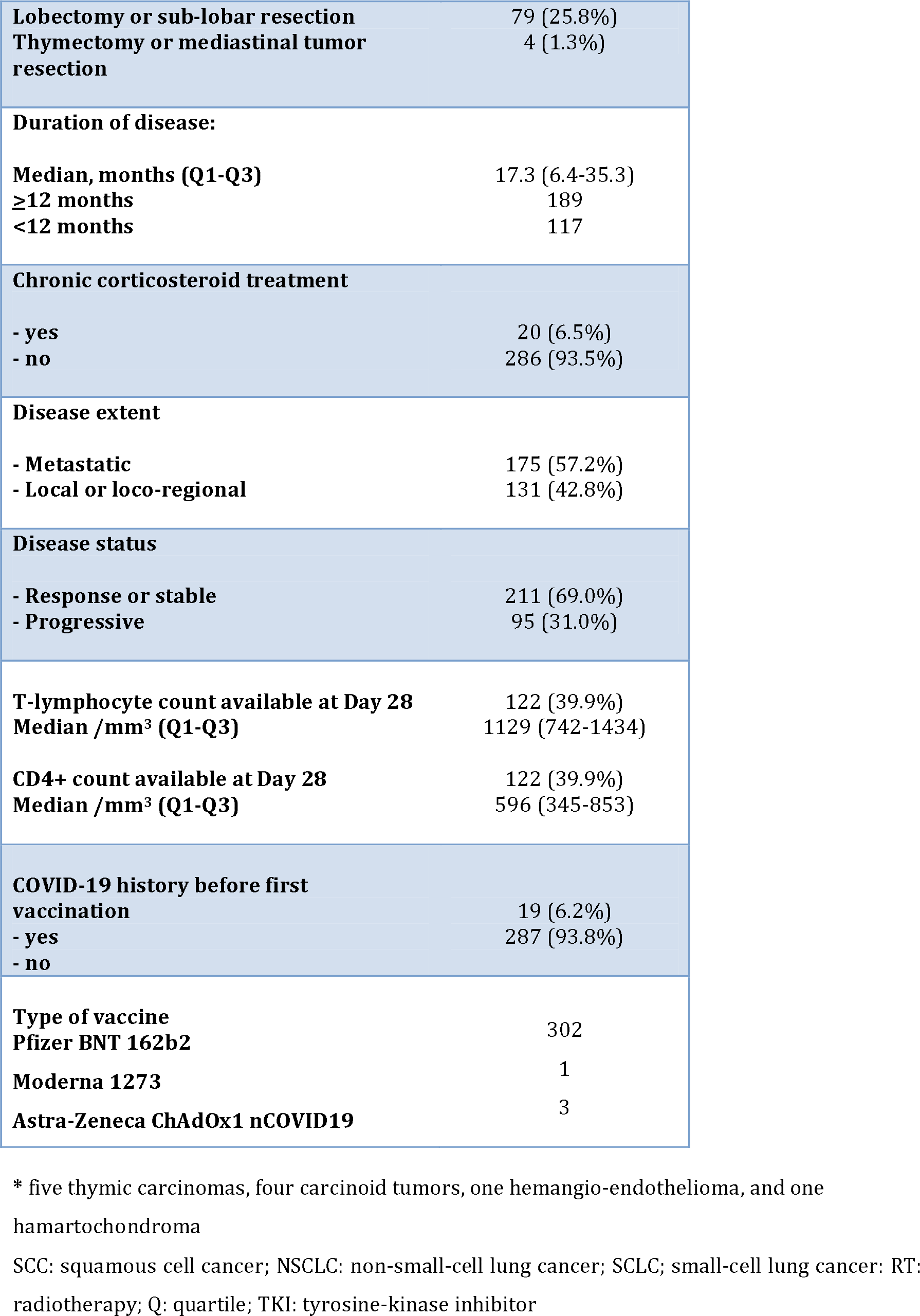
Patient clinical and demographic characteristics at baseline

Total lymphocyte counts were available for 122 (39.9%) patients on Day 28, with a median T-lymphocyte (CD3+) count at 1129/mm^3^ IQR(742-1434), and median CD4+ T-cell count at 596/mm^3^ IQR(345-853).

### Humoral immune response

Median follow-up was 141 days IQR(134-183). Overall, 283 (92.5%) patients underwent serological testing at Day 28 post-first injection and received a second vaccine on that booster day. At Day 28, At Day 28, 248 samples from patients free of prior SARS-CoV-2 infection or with anti-N negative IgG at D0 or D28 (n=265) were available. Median anti-S IgG IgG titer was 149.7 AU/mL IQR(21.9 - 436.1).

In patients without symptomatic or asymptomatic COVID-19 (thus excluding twenty-two patients, 17 with prior COVID-19 history, 3 with PCR-proved COVID-19 post-1st dose, and 2 with anti-N IgG detected at D0) history, a striking increase in antibody titers occurred between Day 0 (137 patients with available serology) and Day 28 (248 patients with available serology) post-first vaccine (**Figure 1A**).

Not all antibodies detected are able to efficiently neutralize the virus by impairing its binding to the ACE2 receptor expressed by respiratory cells. Neutralizing antibodies constitute a variable part of the anti-spike antibodies. The neutralization activity was measured using a pseudo-neutralization assay, assessing neutralizing antibody capacity via competition with ACE2-receptor for binding to anti-spike RBD. **Suppl. Figure 2** depicts the correlation between anti-S IgG log_10_ titers and anti-spike RBD pseudo-neutralization log_10_ titers. A strong correlation was observed from anti-S IgG titer of 300 AU/mL (Spearman’s test, rho= 0.92, p<0.0001), supporting the neutralizing effect of serum anti-S IgG levels levels exceeding such cut-off. At Day 28, 91 patients (32.3%) displayed no anti-S IgG (<50 AU/mL), while 165 patients (58.5%) exhibited only low titers (≤300 AU/mL).

By comparison, the median value of 18 healthy controls was 913 AU/mL IQR(438,3 - 1859.3), antibody titer distribution of healthy controls significantly differing from that of patients treated using chemotherapy (p <0.0001, Mann-Whitney), immunotherapy (p <0.0001, Mann-Whitney), targeted therapy or bevacizumab (p=0.043, Mann-Whitney), or those without systemic therapy <last three months (p <0.0001, Mann-Whitney) (**Figure 1B**). No significant differences in serum anti-spike antibody titers were seen between chemotherapy- and immunotherapy-treated patients (p=0.11).

The second booster dose was not administered to 24 patients due to cancer-related general condition alteration (n=2), mild symptomatic PCR-proven COVID-19 infection post-first vaccine (n=3), history of symptomatic COVID-19 before vaccination supported by anti-N IgG detection (**n=17),** and totally asymptomatic COVID-19as reflected again by anti-N IgG detection (n=2).

Serological data were available for 269 (88%) patients at Day 42, two-to-nine weeks post-second vaccine (median time interval: 52 days IQR(45-69). Of the 306 patients, 37 could not undergo late serological control due to altered general condition (n=4), cancer-related death (n=9), patients refusal (n=5), whereas 19 patients exhibited COVID-19 at any time, with late serological control deemed unnecessary by referent physicians.

Among patients free of prior SARS-CoV-2 infection or with anti-N negative IgG at D0 and D28 (n=265), 248 samples were available at D42 (≥14 days post-second dose) with median serum anti-S IgG titer at 4725 AU/mL IQR(1066 - 13698), 300 AU/mL corresponding to 12.5^th^ percentile.

Two-to-nine weeks post-second vaccine, an overall increase in serum anti-S IgG titers was noted (**Figure 1C**) with a mean 1.4-to-2-fold increase in the log_10_ anti-spike IgG concentrations. However, 17 patients (6.3%) still exhibited negative serological testing, while 34 (11%) displayed IgG concentrations ≤300 AU/mL, with 65 patients (24.1%) exhibiting antibody titers below the first quartile value of 1066 UA/mL. The median serum anti-S IgG concentration in 13 healthy controls, within a median 57-day interval post-second vaccine, was 10594 AU/mL IQR(8350-14836). The titer distribution significantly differed from that observed in patients treated using chemotherapy (p=0.0003, Mann-Whitney), immunotherapy (p=0.013, Mann-Whitney), oral targeted therapy or bevacizumab (p=0.02, Mann-Whitney), or those without systemic therapy <last three months (p=0.001, Mann-Whitney) (**Fig. 1C**). Considering the 231 patients with available data at both points, the anti-S IgG titers significantly rose between Day 28 (first dose) and Day 42 (second dose), irrespective of systemic treatments received (**Figure 2**), with higher titers observed in previously COVID-19-infected patients (n=31) (**suppl. Figure 3**). In patients for whom serology was available at Day 28 post-first (sole) vaccine, an increase by 2 logs in anti-S IgG antibodies was recorded, these antibodies remaining high in 14 patients on later samplings.

**Figure 2:**
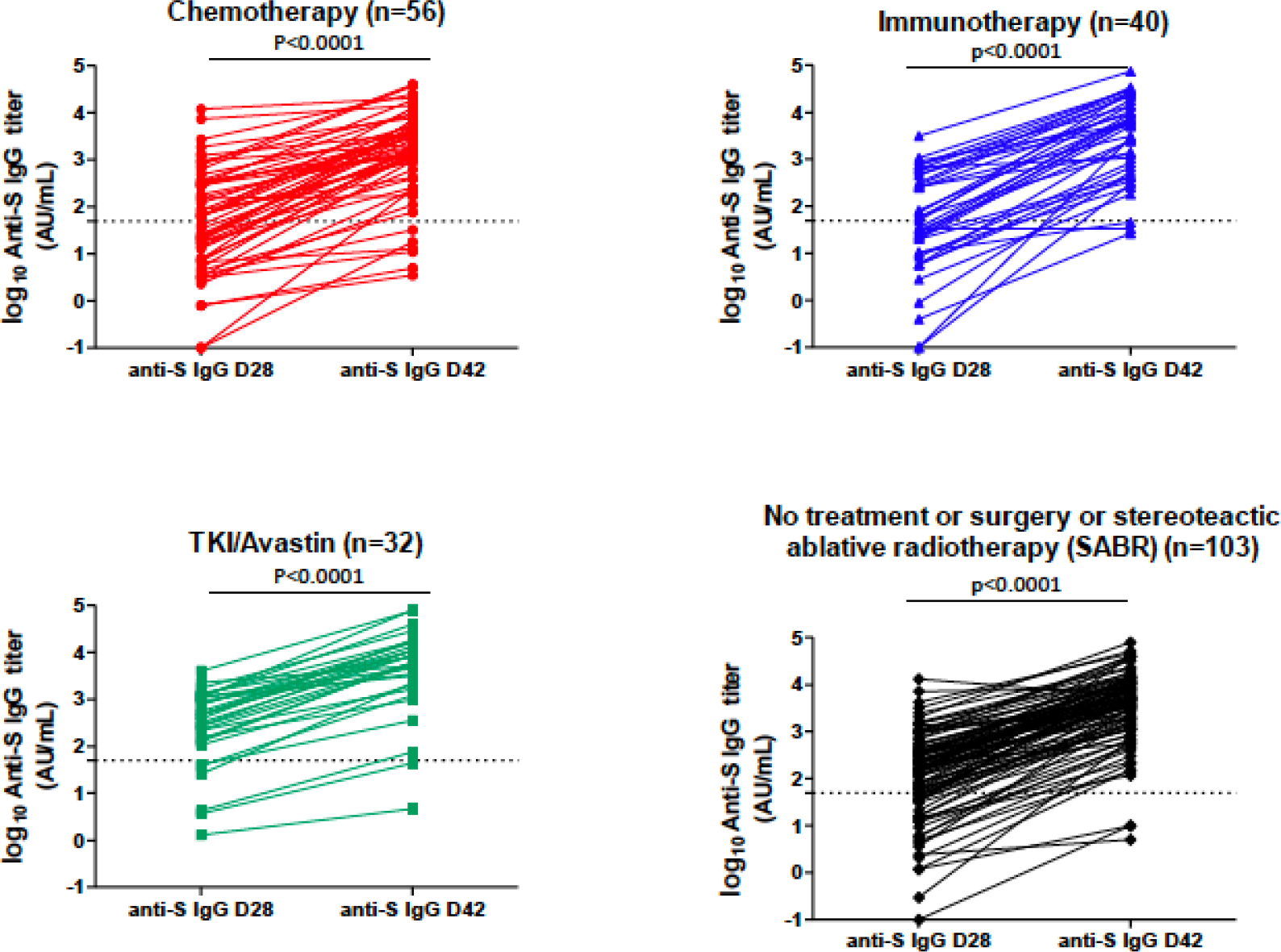
Anti-SARS-Cov-2-spike IgG antibody titers at Day 28 and Day 42 according to the different systemic treatments received. The horizontal dashed lines along the x axes indicate the limit of detection (positivity cut-off) provided by the manufacturer (log_10_ 50 AU/mL). A nonparametric two-tailed pair-wise comparison was performed using the Wilcoxon matched-pairs signed-rank test.

**Figure 3:**
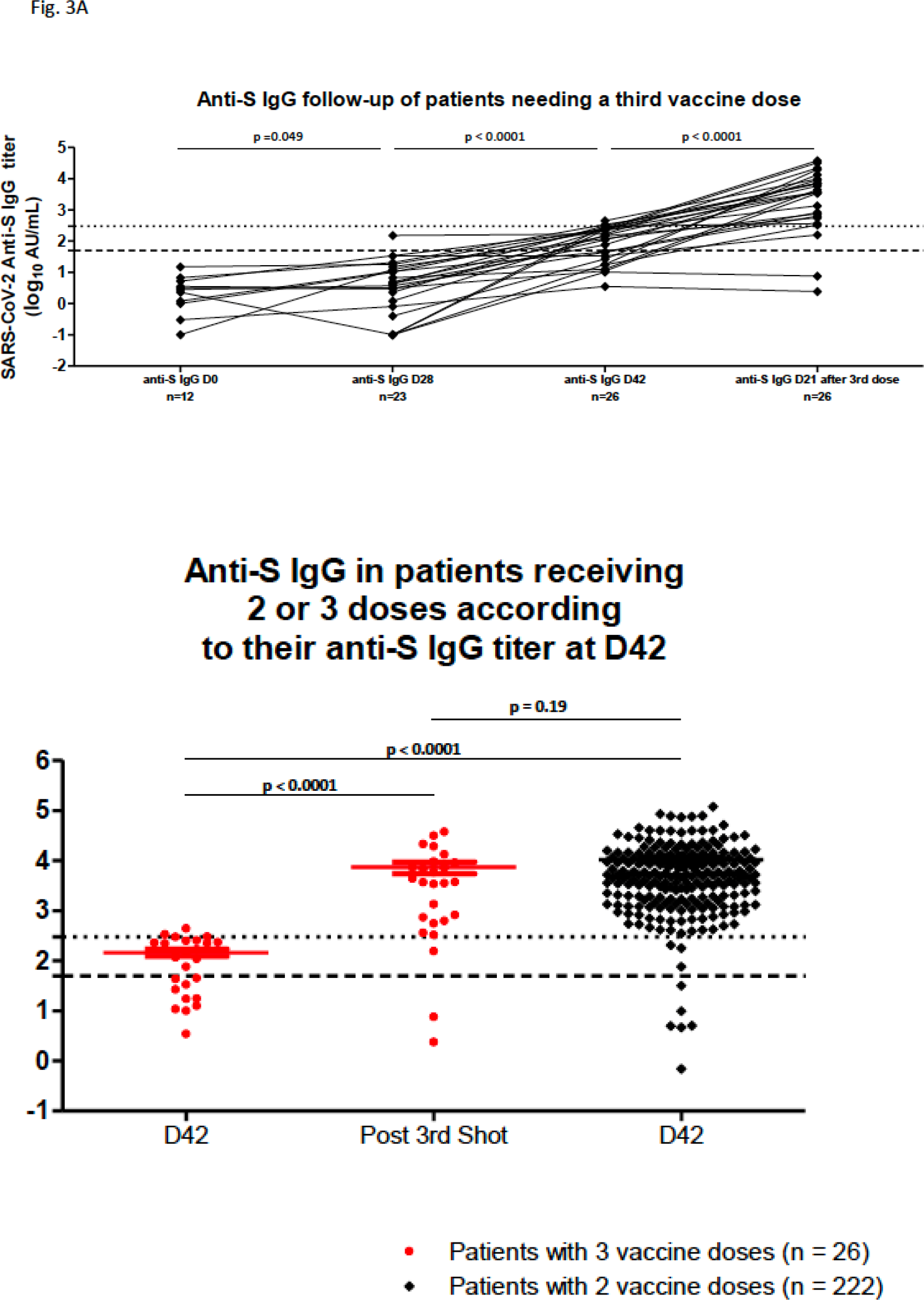
<u>Figure 3A:</u> Evolution from Day 0 of anti-SARS-Cov-2-spike IgG antibody titers after a third vaccine dose injection in 26 patients with titers below 300 AU/mL at post-second dose sampling. The lower dashed line along the x axe indicates the limit of detection (positivity cut-off) provided by the manufacturer (log_10_50 AU/mL). The upper dashed line along the x axe indicates log_10_300 AU/mL cut-off. Statistical comparison used Mann-Whiney test. <u>Figure 3B:</u> Comparison of anti-SARS-Cov-2-spike IgG antibody titers at Day 42 with titers post-third vaccine dose in 26 patients, and with Day 42 titers of 222 patients who received two doses. Statistical comparison used Mann-Whiney test.

During the 6-month follow-up from late January to July 2021, seven patients (2.3%) experienced mildly symptomatic PCR-proven COVID-19 symptoms. In four, these symptoms respectively occurred at Days 4, 6, 12, and 20 post-first vaccine, and in the remaining three, respectively, at Days 33, 35, and 42 post-second vaccine. Only one thymic carcinoma patient (serum anti-S IgG titers at 300.4 AU/mL two days before positive SARS-CoV-2 PCR testing) was hospitalized, due to his frail condition, yet without requiring oxygen supply. He was discharged a week later.

### Safety

No anaphylaxis reaction occurred among the 306 patients, with 587 vaccine doses administered. Safety data were available for 278 patients (90.1%), without significant safety concerns. One-third of patients (n=98) did not report symptoms post-first injection. Reported undesirable effects were transitory pain, injection-site swelling, or Grade 1 injection-site erythema, lasting <24 hours. More frequent undesirable effects of mild intensity were reported post-second vaccination in two-thirds of patients (n=201), including injection-site erythema, pain, local injection-site swelling, mild fever (<38.5°C), all lasting <36 hours. Flu-like symptoms, chills, and fatigue lasting <48 hours were reported in 25 (8%) cases. In one patient, a spectacular Grade 2 urticarial reaction occurred, resolving under oral antihistamines <three days of booster dosing. Another patient reported a large Grade 2 local reaction manifesting within 24 hours post-second injection, as large annular erythema plaques, with pain, fatigue, and mild fever (38°C), with spontaneous resolution within eight days. No vaccine-related death occurred.

### Predictors of lack of immunization

We analyzed the correlation between serological titers using different cut-points (≤50 AU/mL; ≤300 AU/mL) and the main clinical, demographic, and biological variables (**suppl. Table 1A, B, C, D**). At Day 28, age (p<0.0001, **suppl. Fig. 4A**), male gender (p=0.05), chemotherapy-based treatment <three last months (p=0.004), immunotherapy as single-therapy <three last months (p=0.01), long-term corticosteroid treatment (p=0.01), and lack of disease control (p=0.03), were significantly associated with negative serological testing ≤50 AU/mL in univariable analyses (**suppl. Table 1A**). Conversely, each 100 units/mm3 increase in Day 28 T-lymphocyte (CD3+) counts (p=0.008), and Day 28 CD4+ T-cell counts (p=0.002, **suppl. Fig. 4B**), were associated with higher seroconversion probability (n=122). In 111 patients for whom these analyses were performed, Day 28 interferon-γ specific T-cell response to SARS-CO-2-spike measured by ELISPOT assay was significantly associated with higher seroconversion probability at Day 28 (OR=0.968; 95%CI: 0.941-0.995, p=0.021).

Considering variables exhibiting a p=0.2 value in univariable analyses, except for the immunological data (n=122 or 111), multivariable logistic regression analyses (**suppl. Table 1A**) confirmed that age (adjusted OR=1.05; 95%CI :1.02-1.08, p=0.0001), with a 5% increase in non-immunization risk at Day 28 for each year, long-term corticosteroids (adjusted OR=3.26; 95%CI:1.1-9.6, p=0.033), and chemotherapy as last treatment <three last months (adjusted OR=3.0; 95%CI: 1.05-8.4, p=0.041) were independently associated with antibody titers ≤50 AU/mL at Day 28.

Considering Day 28 cut-off at ≤300 AU/mL (**suppl. Table 1B**), age (p<0.0001, suppl. Fig. 4C), male gender (p=0.019), chemotherapy (p=0.007), or immunotherapy (p=0.037) as last treatment were significantly associated with anti-S IgG titers ≤300 AU/mL. Although T-lymphocyte and CD4+ T-cell counts failed to predict seroconversion at this cut-off, Day 28 interferon-γ specific T-cell responses to SARS-CO-2-spike (ELISPOT assay; n=90), was highly predictive of anti-S IgG titers over 300 AU/mL (OR=0.956; 95% IC: 0.935-0.978, p<0.0001) in 111 patients with available results. In multivariable analyses, only age, (adjusted OR= 1.05, 95%CI:1.02-1.7, p <0.0001) and chemotherapy as last treatment received (adjusted OR=3.07; 95%CI:1.36-6.94, p=0.048) were independently associated with Day 28 anti-S IgG titers ≤300 AU/mL.

Considering Day 42 cut-off at ≤50 AU/mL (post-second vaccination), the variables significantly associated with negative serology risk included age (p=0.013), lack of disease control (p=0.015), chronic corticosteroids (p=0.007), and chemotherapy as last treatment received (p=0.021) (**suppl. Table 1C**). Conversely, in 74 patients with available data, each 100 units/mm3 increase in Day 42 T-cell (OR= 0.60; 95%CI:0.40-0.89, p=0.012) or CD4+ T-cell counts (OR=0.15; 95%CI: 0.35-0.63, p=0.009) was associated with a higher seroconversion probability. In multivariable analyses, age (adjusted OR=1.09; 95%CI:1.03-1.15, p=0.004), chemotherapy as last treatment received (adjusted OR= 6.60; 95%CI:1.05-41.62, p=0.045), and long-term corticosteroids (adjusted OR= 6.22; 95%CI:1.41-27.43, p=0.016), were significantly associated with negative Day 42 serology (≤50 AU/mL).

Considering Day 42 cut-off at ≤300 AU/mL (**suppl. Table 1D**), age (p=0.028) metastatic disease extent (p=0.014), chemotherapy <three last months (p= 0.004) long-term corticosteroids (p< 0.00001), and lack of disease control (p=0.010) were associated with lower immunization probability. Conversely, cancer duration was associated with a higher Day 42 sero-conversion probability >300 AU/mL (p=0.01). In 74 patients with immunological data, each 100 units/mm^3^ increase in Day 42 T-cell (OR=0.73; 95%CI:0.59-0.90, p=0.003) or CD4+ T-cell counts (OR=0.53; 95%CI:0.35-0.82, p=0.005) was associated with higher seroconversion probability. Only age (adjusted OR=1.07; 95%CI:1.02-1.11, p=0.002), chemotherapy as last treatment received (adjusted OR=3.14; 95%CI:1.08-9.13, p=0.036), and long-term corticosteroids (adjusted OR=6.2, 95%CI:1.4-27.4, p=0.016) independently influenced the probability of low response (anti-SARS-COV-2-spike IgG titers ≤300 AU/mL) in multivariable analyses, diminishing the probability of a reliable protection against SARS-CoV-2 infection.

### Third vaccine dose

Serial serological tests were performed in ten patients exhibiting low antibody titers post-vaccine boosting (≤300 AU/mL). In seven, the antibody titers decreased over time (n=6) or remained stable (n=1), within a 13-52 day period, whereas the three others displayed a slight increase over 300 AU/mL, within 59 days post-second dose. Two patients still presented <1066 AU/mL, 53 and 47 days post-booster dose, remaining <25^th^ percentile. Seven of these patients were still receiving chemotherapy-based treatment (n=5) or corticosteroids (n=2), with three receiving neither chemotherapy nor corticosteroids.

At Day 42, 30 patients exhibiting anti-S IgG titers ≤300 AU/mL were proposed a third vaccine, from Day 28 post-second shot. Of these, two experienced cancer-related condition deterioration; therefore, they underwent no serology control post-third injection. A serology assay was available beyond Day 21 post-third injection in 26/30 remaining (results still awaited for two). At the time of analysis, none of these patients displayed symptomatic COVID-19. In 26 (86.7%) patients with serological tests available at Day 28 post-third shot (**Figure 3A**), 19 (73%) demonstrated a dramatic rise in anti-S IgG titers, exceeding 3500 AU/mL, four (15.4%) displayed a moderate increase beyond 300 AU/mL cut-point, yet <1000 AU/mL (**Figure 3B**). Therefore, 88.5% exhibited sero-conversion. In all, persistent negative anti-N IgG were found, excluding any recent SARS-CoV-2 infection. Among these, three had been receiving corticosteroids for several weeks, which were continued at third vaccination. Only three patients did not respond to third vaccination, all being older than 65 years, being either totally negative (n=2, <50/AU/mL) or exhibiting low anti-spike antibodies. Among them was patient (still <50 AU/mL post-second booster), on chronic myelomonocytic leukemia Type 2, with complete molecular response. The two other patients, respectively displaying 47 and 157 AU/mL, were treated using either chemo-immunotherapy or ICI alone. These two latter patients, also suffered from hematological conditions (hypogammaglobulinemia, monoclonal IgG peak), possibly explaining their poor immunization.

## DISCUSSION

COVID-19 vaccines were made available in France in January 2021. Nothing was known on COVID-19 vaccination efficacy in patients with poor immune conditions, including metastatic lung cancer patients, especially those under systemic corticosteroids or cytotoxic chemotherapy. Though lung cancer patients were reported at high COVID-19-related mortality risk in published series, lethality systematically exceeding 30% of infected patients^1, 3, 28^, we observed only seven mild COVID-19 cases among our 306 vaccinated patients (2.3%). Such observation strongly supports the efficacy of mRNA COVID-19 vaccines used in 98.4% of our population. A possible limitation to such outcome is the decrease in SARS-CoV-2 virus circulation <0.79 in France by mid-May 2021, versus 1.2 or 1.3 in late January 2021. However, a dramatic rise in infections occurred early July, resulting in >20,000 new daily cases. The patient acceptation rate of systematic vaccination was in line with previous reports, with only 11% initial refusals^29^. Reactogenicity was weak, without short-term serious adverse effects in this real-life setting. We did not observe specific safety concerns in ICI-treated patients, especially regarding immune-related side-effects, as reported by Israeli teams^30^. Moreover, our study emphasized that sero-conversion monitoring could be useful in immuno-suppressed patients. In this population, the first vaccine efficacy was much lower than that reported in vaccine registration trials, with one-third of patients displaying negative serological testing (≤50 AU/mL) at Day 28, whereas three-quarters exhibited <25^th^ percentile serological titer distribution. These data are in line with prospective studies involving a mixed population with solid cancers and hematological malignancies^31, 32^. Although there has been no clear cut-off for antibody titers predicting protection against severe COVID-19, a 300 AU/mL cut-off was shown to well correlate with the pseudo-neutralization assay, as a readout for anti-viral efficacy. We thus selected this value as protection cut-off against SARS-CoV-2 infection in our patients^33^. Let us keep in mind that the recently described delta strain, which currently represents more than 90% of sequenced viral isolates in France^34^, was reported to be 40-80% more transmissible than the alpha strain^35, 36^, its viral burden being 1000 fold higher than other strains^37^. It is thus crucial to define serological correlates confirming the protection of immuno-compromised patients.

With this in mind, our study provided strong evidence for keeping the initially established intervals between two vaccine shots for cancer patients. These patients displayed a delay in their immunization process, with lower levels of protective circulating vaccination-induced antibodies versus healthy vaccinated controls. Conversely, a reassuring observation has been the booster injection’s remarkable efficacy, with only 6.0% of thoracic cancer patients still displaying negative serology at Day 42, whereas only % exhibited antibody titers ≤300 AU/mL. The two characteristics independently associated with poor immunization, irrespective the cut-point chosen, were age and long-term corticosteroids. Concerning age, a lower immunization rate was identified in octogenarians, along with a 5% decreased probability per year to reach protective immunization. Regarding long-term corticosteroids, the lower immunization may probably be explained by either lower total T-cell and CD4+ T-lymphocyte counts or T-cell specific responses to spike protein. The adverse impact of age on the ability to induce vaccine-related protective humoral responses was already highlighted in a study involving octogenarians^38, 39^. While clearly delaying the immunization process as previously reported^31, 40, 41^, cytotoxic chemotherapy was associated with higher low-immunization rates at Day 42, as well.

As herein shown, a third vaccine could contribute to appropriate sero-protection in patients still poorly immunized post-two vaccines. Overall, 92% of patients were shown to benefit from a third shot, reflected by substantial increases in anti-S IgG antibodies, with only very few patients left without sufficient protection. A limitation to this observation is the group’s small sample size. Such outcome must be confirmed in larger-scale studies involving solid cancer patients. Notably, a third vaccine dose is still being debated in patients with hematological malignancies or solid organ transplantation^42–47^. This latter statement is supported by the fact that three of our patients with negative serology post-third vaccine were, indeed, suffering from underlying hematological conditions. Consequently, patients with lymphocyte function defects due to lymphoid cancers or lymphocyte-depleting treatments^44^ may exhibit lower benefits from a third vaccination.

## Data Availability

All the data are available on request to the senior authoor of the article, Prof. Gerard ZALCMAN

## List of supplementary material

**Supplemental Figure 1:**
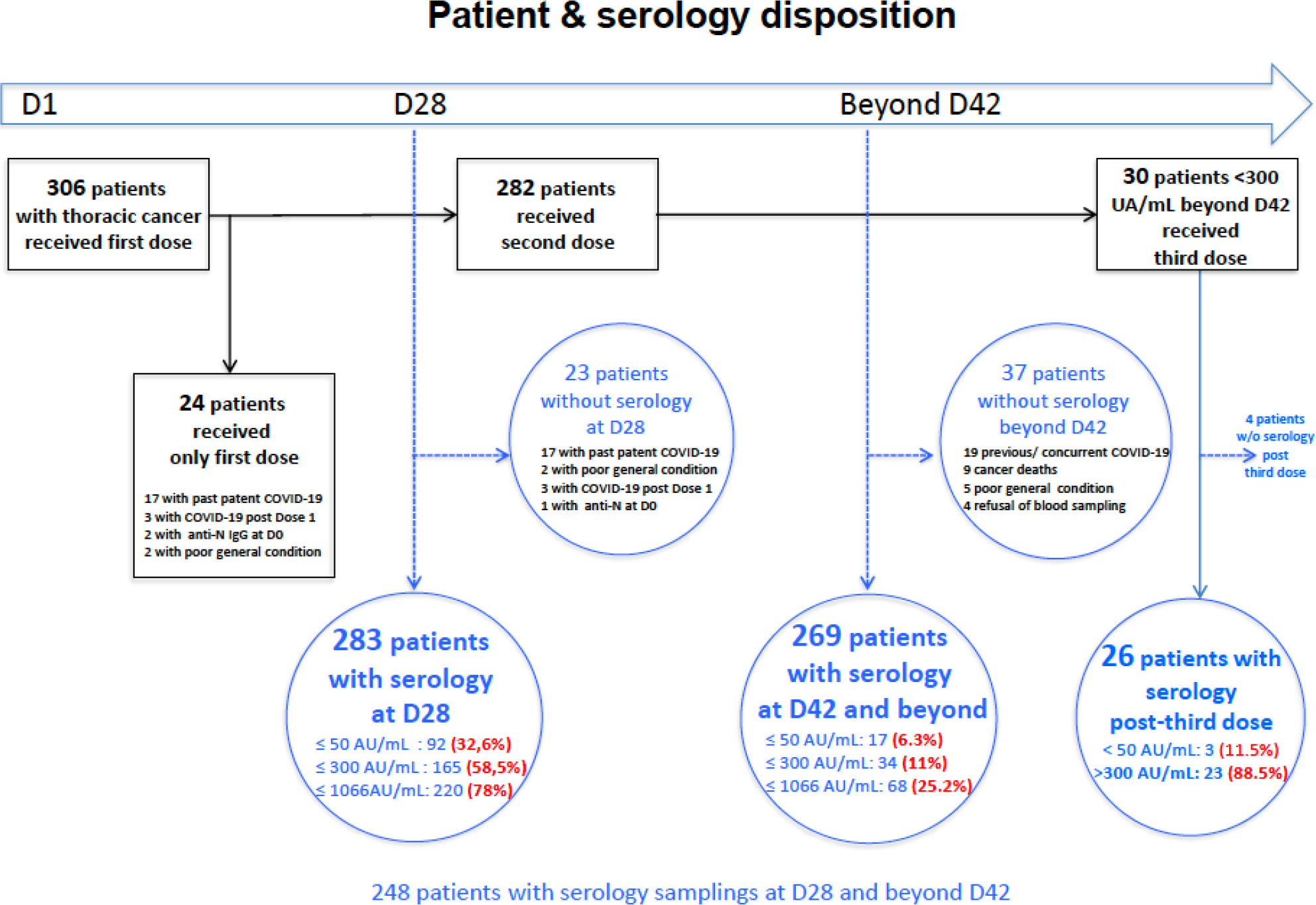
Patients disposition

**Supplemental Figure 2:**
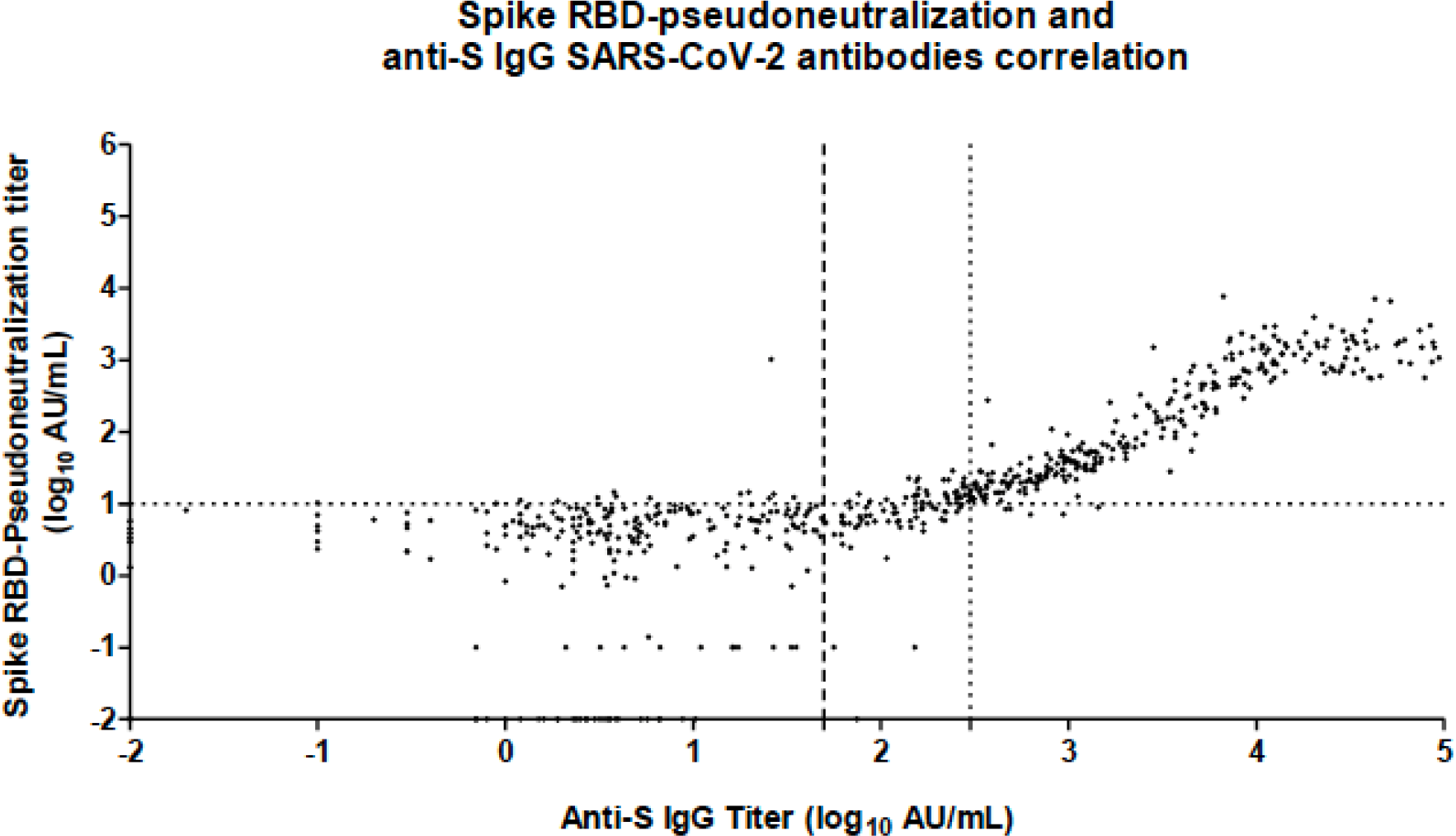
Correlation between anti-SARS-Cov-2-spike IgG antibody titers (Log_10_ Arbitrary Unit/mL), x axis, and -spike RBD-pseudo-neutralization titers (Log_10_ AU/mL), y axis. Vertical dashed lines correspond to the 50 AU/mL (cut-point for positivity according to the manufacturer) and 300 AU/mL anti-SARS-Cov-2-spike IgG antibody log_10_ titers. The two assays showed an excellent correlation (Spearman’s test, rho= 0.92, p<0.0001).

**Supplemental Figure 3:**
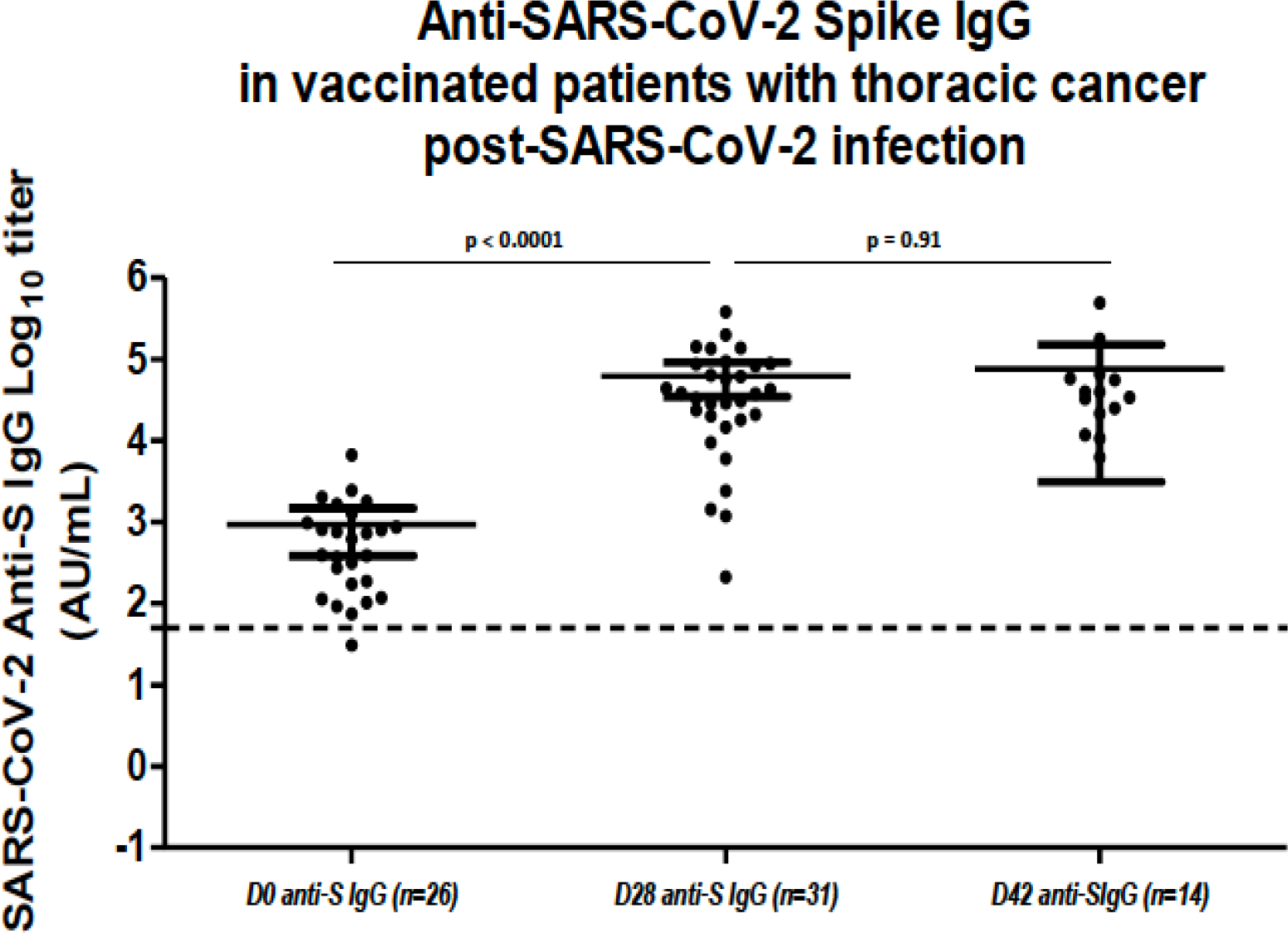
Anti-SARS-Cov-2-spike IgG antibody titers, at Day 0, Day 28, and Day 42, (respectively n=26, n=31, and n=14) in 34 patients with previous COVID-19-history or positive anti-SARS-Cov-2-Nucleocapsid IgG serology. The horizontal dashed line along the x axe indicates the limit of detection (positivity cut-off) provided by the manufacturer (log10 50 AU/mL). Statistical comparison used Mann-Whiney test.

**Supplemental Figure 4A, 4B & 4C:**
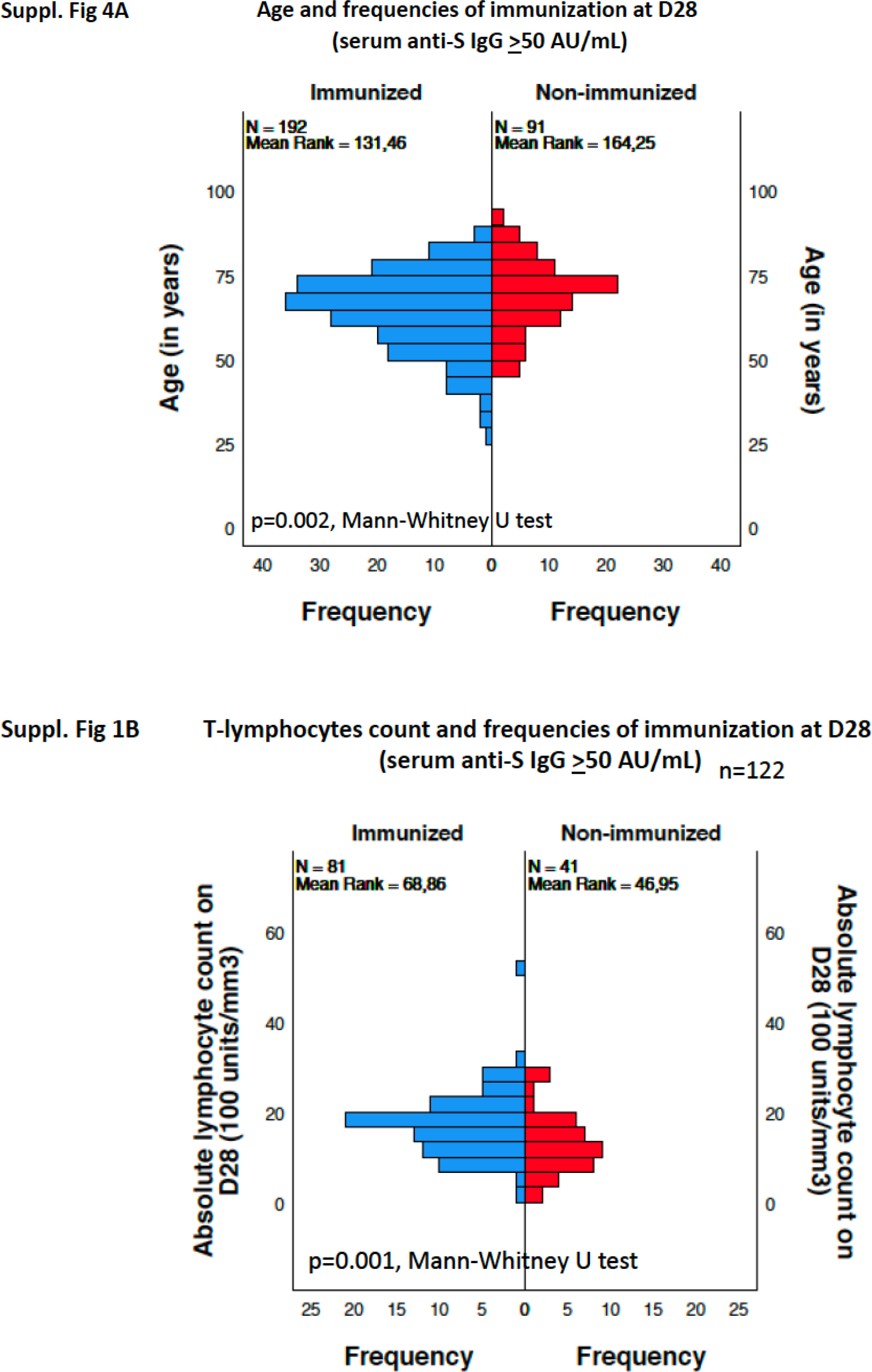
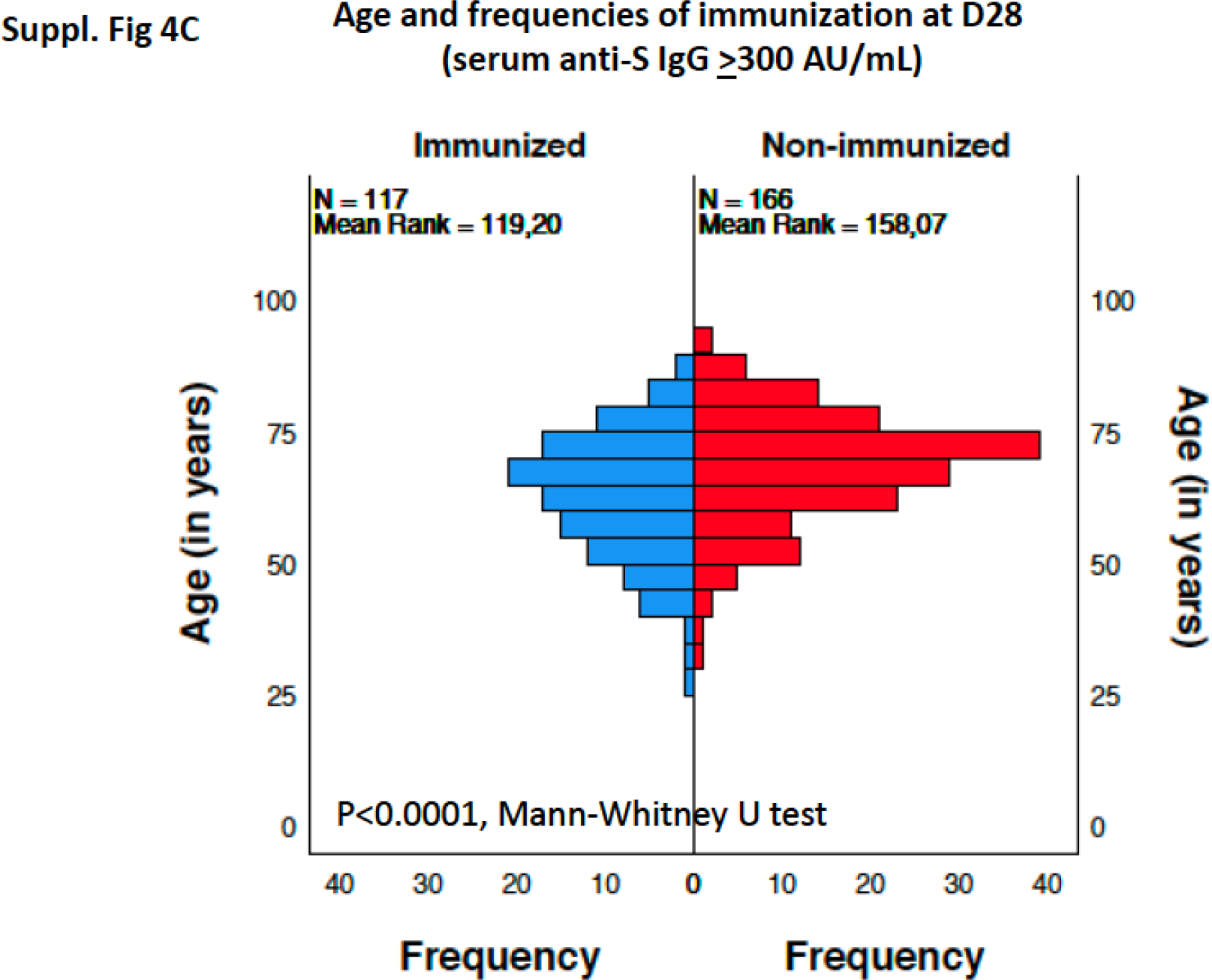
<u>Suppl. Figure 4A:</u> Distribution of patient ages according to Day 28 anti-S IgG antibody titers, above or below 50 AU/mL. Statistical comparison used Mann-Whiney U test. <u>Suppl. Figure 4B:</u> Distribution of patients' T-lymphocyte counts according to Day 28 anti-S IgG antibody titers, above or below 50 AU/mL. Statistical comparison used Mann-Whiney U test. <u>Suppl. Figure 4C:</u> Distribution of patient ages according to Day 28 anti-S IgG antibody titers, above or below 300 UA/mL. Statistical comparison used Mann-Whiney U test.

## Supplemental Immunology methods

**Supplemental Table 1A:**
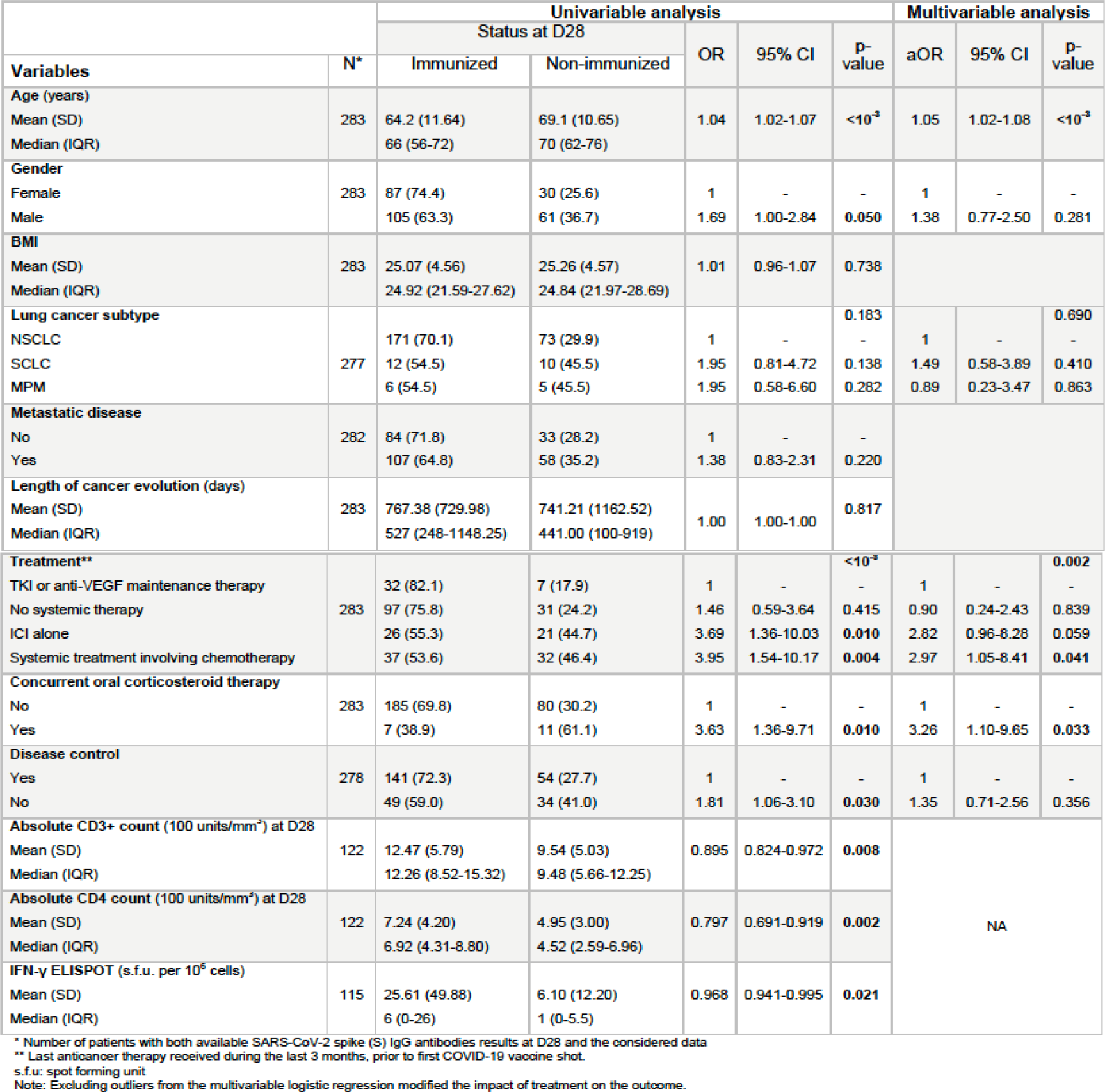
Predictors of non-immunization (anti-S IgG antibodies at 50 AU/mL cut-off) at D28

**Supplemental Table 1B:** Predictors of non-immunization (anti-S IgG antibodies at 300 AU/mL cut-off) at D28

| Variables | N* | Univariable analysis |  |  |  |  | Multivariable analysis |  |  |
| --- | --- | --- | --- | --- | --- | --- | --- | --- | --- |
|  |  | Status at D28 |  | OR | 95% CI | p-value | aOR | 95% CI | p-value |
|  |  | Immunized | Non-immunized |  |  |  |  |  |  |
| **Age (years)** |  |  |  |  |  |  |  |  |  |
| Mean (SD) | 283 | 62.6 (11.82) | 68 (10.82) | 1.04 | 1.02-1.07 | <10<sup>-3</sup> | 1.05 | 1.02-1.07 | <10<sup>-3</sup> |
| Median (IQR) |  | 64 (54-70) | 69 (61-75) |  |  |  |  |  |  |
| **Gender** |  |  |  |  |  |  |  |  |  |
| Female | 283 | 58 (49.6) | 59 (50.4) | 1 | - | - | 1.58 | 0.93-2.70 | 0.089 |
| Male |  | 59 (35.5) | 107 (64.5) | 1.78 | 1.10-2.89 | 0.019 |  |  |  |
| **BMI** |  |  |  |  |  |  |  |  |  |
| Mean (SD) | 283 | 25.19 (4.59) | 25.09 (4.54) | 0.99 | 0.95-1.05 | 0.855 |  |  |  |
| Median (IQR) |  | 25.07 (21.85-27.18) | 24.78 (21.90-28.43) |  |  |  |  |  |  |
| **Lung cancer subtype** |  |  |  |  |  | 0.054 |  |  | 0.204 |
| NSCLC |  | 107 (43.9) | 137 (56.1) | 1 | - | - | 1 | - | - |
| MPM | 277 | 3 (27.3) | 8 (72.7) | 2.08 | 1.16-10.69 | 0.287 | 1.28 | 0.31-5.32 | 0.731 |
| SCLC |  | 4 (18.2) | 18 (81.8) | 3.52 | 0.54-8.04 | 0.027 | 2.81 | 0.89-8.86 | 0.077 |
| **Metastatic disease** |  |  |  |  |  |  |  |  |  |
| Yes | 282 | 68 (41.2) | 97 (58.8) | 1 | - | - |  |  |  |
| No |  | 48 (41.0) | 69 (59.0) | 1.01 | 0.62-1.63 | 0.975 |  |  |  |
| **Length of cancer evolution (days)** |  |  |  |  |  |  |  |  |  |
| Mean (SD) | 283 | 772.66 (634.14) | 749.31 (1035.04) | 1.00 | 1.00-1.00 | 0.828 |  |  |  |
| Median (IQR) |  | 599 (286-1195) | 475.5 (133.75-938.5) |  |  |  |  |  |  |
| **Treatment\*\*** |  |  |  |  |  | 0.047 |  |  | 0.187 |
| TKI or anti-VEGF maintenance therapy |  | 23 (59.0) | 16 (41.0) | 1 | - | - | 1 | - | - |
| No systemic therapy | 283 | 55 (43.0) | 73 (57.0) | 1.91 | 0.92-3.95 | 0.082 | 1.41 | 0.63-3.13 | 0.403 |
| ICI alone |  | 17 (36.2) | 30 (63.8) | 2.54 | 1.06-6.07 | 0.037 | 1.94 | 0.76-4.97 | 0.165 |
| Systemic treatment involving chemotherapy |  | 22 (31.9) | 47 (68.1) | 3.07 | 1.36-6.94 | 0.007 | 3.07 | 1.36-6.94 | 0.048 |
| **Concurrent oral corticosteroid therapy** |  |  |  |  |  |  |  |  |  |
| No | 283 | 113 (42.6) | 152 (57.4) | 1 | - | - | 1 | - | - |
| Yes |  | 4 (22.2) | 14 (77.8) | 2.60 | 0.83-8.11 | 0.099 | 2.07 | 0.63-6.86 | 0.232 |
| **Disease control** |  |  |  |  |  |  |  |  |  |
| Yes | 278 | 31 (37.3) | 52 (62.7) | 1 | - | - |  |  |  |
| No |  | 86 (44.1) | 109 (55.9) | 1.32 | 0.78-2.24 | 0.297 |  |  |  |
| **CD3+ T-cell count (100 units/mm<sup>3</sup>) at D28** |  |  |  |  |  |  |  |  |  |
| Mean (SD) | 122 | 12.23 (6.23) | 11.13 (5.43) | 0.967 | 0.905-1.034 | 0.326 |  |  |  |
| Median (IQR) |  | 11.30 (8.35-15.16) | 11.11 (6.45-14.05) |  |  |  |  |  |  |
| **CD4+ T-cell count (100 units/mm<sup>3</sup>) at D28** |  |  |  |  |  |  |  |  |  |
| Mean (SD) | 122 | 7.35 (4.92) | 6.06 (3.40) | 0.922 | 0.834-1.020 | 0.116 |  |  |  |
| Median (IQR) |  | 6.92 (4.32-8.56) | 5.33 (3.18-9.43) |  |  |  |  |  |  |
| **IFN-γ ELISPOT (s.f.c. per 10<sup>6</sup> cells)** |  |  |  |  |  |  |  |  |  |
| Mean (SD) | 115 | 45.89 (66.62) | 6.99 (12.38) | 0.956 | 0.935-0.978 | <10<sup>-3</sup> |  |  |  |
| Median (IQR) |  | 18 (4-72) | 1 (0-8) |  |  |  |  |  |  |

\* Number of patients with both available SARS-CoV-2 spike (S) IgG antibodies results at D28 and the considered data
\*\* Last anticancer therapy received during the last 3 months, prior to first COVID-19 vaccine shot
s.f.u.: spot forming unit
Note: Excluding outliers from the multivariable logistic regression modified the impact of treatment on the outcome.

**Supplemental Table 1C:**
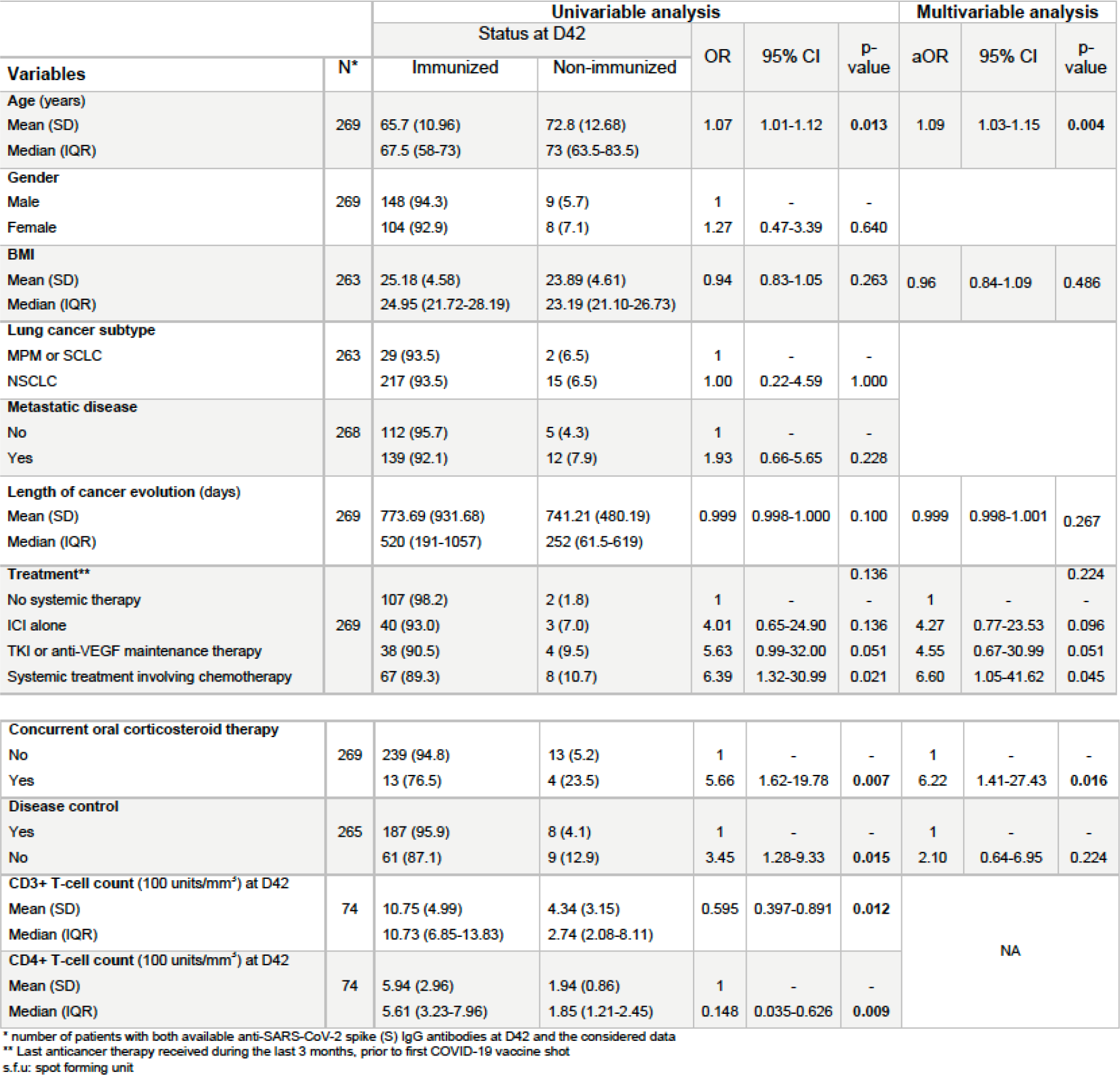
Predictors of non-immunization (anti-S IgG antibodies at 50 AU/mL cut-off) at D42

**Supplemental Table 1D:** Predictors of non-immunization (anti-S IgG antibodies at 300 AU/mL cut-off) at D42

| Variables | N* | Univariable analysis |  |  |  |  | Multivariable analysis |  |  |
| --- | --- | --- | --- | --- | --- | --- | --- | --- | --- |
|  |  | Status at D42 |  | OR | 95% CI | p-value | aOR | 95% CI | p-value |
|  |  | Immunized | Non-immunized |  |  |  |  |  |  |
| **Age (years)** |  |  |  |  |  |  |  |  |  |
| Mean (SD) | 269 | 65.6 (10.98) | 70.2 (11.93) | 1.04 | 1.00-1.08 | 0.028 | 1.07 | 1.02-1.11 | 0.002 |
| Median (IQR) |  | 68 (58-73) | 70 (61.5-81.25) |  |  |  |  |  |  |
| **Gender** |  |  |  |  |  |  |  |  |  |
| Female | 269 | 99 (88.4) | 13 (11.6) | 1 | - | - |  |  |  |
| Male |  | 136 (86.6) | 21 (13.4) | 1.18 | 0.56-2.46 | 0.667 |  |  |  |
| **BMI** |  |  |  |  |  |  |  |  |  |
| Mean (SD) | 269 | 25.26 (4.65) | 23.97 (3.91) | 0.94 | 0.86-1.02 | 0.124 | 0.97 | 0.88-1.07 | 0.513 |
| Median (IQR) |  | 24.97 (21.72-28.44) | 23.67 (21.38-26.96) |  |  |  |  |  |  |
| **Lung cancer subtype** |  |  |  |  |  |  |  |  |  |
| NSCLC | 263 | 204 (87.9) | 28 (12.1) | 1 | - | - |  |  |  |
| MPM or SCLC |  | 25 (80.6) | 6 (19.4) | 1.75 | 0.66-4.63 | 0.261 |  |  |  |
| **Metastatic disease** |  |  |  |  |  |  |  |  |  |
| No | 268 | 109 (93.2) | 8 (6.8) | 1 | - | - | 1 | - | - |
| Yes |  | 125 (82.8) | 26 (17.2) | 2.83 | 1.23-6.52 | 0.014 | 2.13 | 0.82-5.51 | 0.119 |
| **Length of cancer evolution (days)** |  |  |  |  |  |  |  |  |  |
| Mean (SD) | 269 | 801.21 (950.09) | 413.56 (483.37) | 0.999 | 0.998-1.000 | 0.010 | 0.999 | 0.998-1.000 | 0.081 |
| Median (IQR) |  | 534 (219-1088) | 194 (51.5-602) |  |  |  |  |  |  |
| **Treatment\*\*** |  |  |  |  |  | 0.039 |  |  | 0.209 |
| No systemic therapy |  | 117 (92.9) | 9 (7.1) | 1 | - | - | 1 | - | - |
| TKI or anti-VEGF maintenance therapy | 269 | 32 (86.5) | 5 (13.5) | 2.03 | 0.64-6.49 | 0.232 | 2.12 | 0.55-8.20 | 0.276 |
| ICI alone |  | 38 (86.4) | 6 (13.6) | 2.05 | 0.69-6.14 | 0.198 | 2.33 | 0.66-8.20 | 0.187 |
| Systemic treatment involving chemotherapy |  | 48 (77.4) | 14 (22.6) | 3.79 | 1.54-9.35 | 0.004 | 3.14 | 1.08-9.13 | 0.036 |
| **Concurrent oral corticosteroid therapy** |  |  |  |  |  |  |  |  |  |
| No | 269 | 226 (89.7) | 26 (10.3) | 1 | - | - | 1 | - | - |
| Yes |  | 9 (52.9) | 8 (47.1) | 7.73 | 2.74-21.76 | <10<sup>-4</sup> | 6.54 | 1.95-21.93 | 0.002 |
| **Disease control** |  |  |  |  |  |  |  |  |  |
| Yes | 265 | 177 (90.8) | 18 (9.2) | 1 | - | - | 1 | - | - |
| No |  | 55 (78.6) | 15 (21.4) | 2.68 | 1.27-5.67 | 0.010 | 1.49 | 0.62-3.61 | 0.376 |
| **CD3+ T-cell count (100 units/mm<sup>3</sup>) at D42** |  |  |  |  |  |  |  |  |  |
| Mean (SD) | 74 | 11.08 (5.04) | 5.86 (3.36) | 0.730 | 0.592-0.900 | 0.003 | NA |  |  |
| Median (IQR) |  | 11.12 (7.24-14.41) | 5.85 (2.50-7.70) |  |  |  |  |  |  |
| **CD4+ T-cell count (100 units/mm<sup>3</sup>)** |  |  |  |  |  |  |  |  |  |
| Mean (SD) | 74 | 6.11 (2.97) | 3.07 (2.03) | 0.533 | 0.345-0.823 | 0.005 |  |  |  |
| Median (IQR) |  | 5.73 (3.82-8.11) | 3.00 (1.78-3.43) |  |  |  |  |  |  |

\* number of patients with both available anti-SARS-CoV-2 spike (S) IgG antibodies at D42 and the considered data
\*\* Last anticancer therapy received during the last 3 months, prior to first COVID-19 vaccine shot

## Supplemental online Materials

### Peripheral blood mononuclear cells

Peripheral blood mononuclear cells (PBMCs) were isolated by density gradient centrifugation using Lymphosep™ (1.077 g/ml, Biosera, France). 6 ml of whole blood was diluted 1:1 with PBS. 15 ml of Lymphosep was pipetted into 50-ml SepMate™ centrifuge tubes (Stem Cell Technologies), and the diluted blood was layered over the Lymphosep gradient. The tubes were centrifuged for 10 min at 1200 × g with break. After centrifugation, PBMCs were collected and washed twice in PBS. For cryopreservation, PBMCs were diluted with 4°C heat-inactivated fetal calf serum (FCS, Gibco™, Thermo Fischer Scientific) containing 10% DMSO and stored at −80°C until further use.

The cryopreserved PBMC samples were thawed in a 37°C water bath with gently agitation. Thereafter, the cell suspension was resuspended in 10 ml of pre-warmed RPMI 1640 medium (Gibco™, Thermo Fischer Scientific) supplemented with 10% heat-inactivated FCS and 100 UI/ml Penicillin/Strepomycin (Gibco™, Thermo Fischer Scientific). After centrifugation, cells were resuspended in complete RPMI medium and counted using a hematology analyzer XE-5000 (Sysmex, Kobe, Japan).

### ELISpot assay

T cell responses to SARS-CoV-2 vaccination were assessed by IFN-γ ELISpot assay using a T-SPOT.COVID kit (Oxford Immunotec, Abingdon, UK). Thawed PBMCs were resuspended at 2.5x10^6^ lymphocytes/ml in AIMV medium (Thermo Fischer Scientific). PBMCs were seeded (100 μl per well) and stimulated for 16-18 h with a pool of SARS-CoV-2 spike peptides at 37⍰°C, 5% CO_2_, 95% humidity. Subsequently, cells were washed off and released IFN-γ was detected following the manufacturer’s instructions. Spot forming units (SFU) were counted manually using a DX-1 Microscope (Veho, UK). Results were reported as SFU per million lymphocytes. The unspecific background (SFU from negative control wells) was subtracted from experimental readings. We excluded the results if the positive control well (phytohemagglutinin) was negative. The lower limit to indicate a positive response was 32 spots per million lymphocytes.

### Enumeration of peripheral blood lymphocytes

T-(CD3+, CD3+CD4+, CD3+CD8+), B-(CD19+), and Natural Killer (NK)-lymphocyte (CD3negCD16 + CD56+) absolute counts were enumerated by multiparametric flow cytometry of fresh EDTA-anticoagulated whole peripheral blood (PB). PB samples were processed using a BD FACSDuet™ preparation system integrated with a BD FACSLyric™ flow cytometer (Becton Dickinson Biosciences, San Jose, CA). Lymphocyte subpopulations were assessed using BD Multitest™ CD3/CD8/CD45/CD4, BD Multitest™ CD3/CD16+CD56/CD45/CD19 and BD Trucount™ tubes for absolute count with a single step “lyse-no-wash” procedure (BD FACS™Lysing Solution). BD FACS Suite™ Clinical software version 1.4 was used to collect and analyze the data.

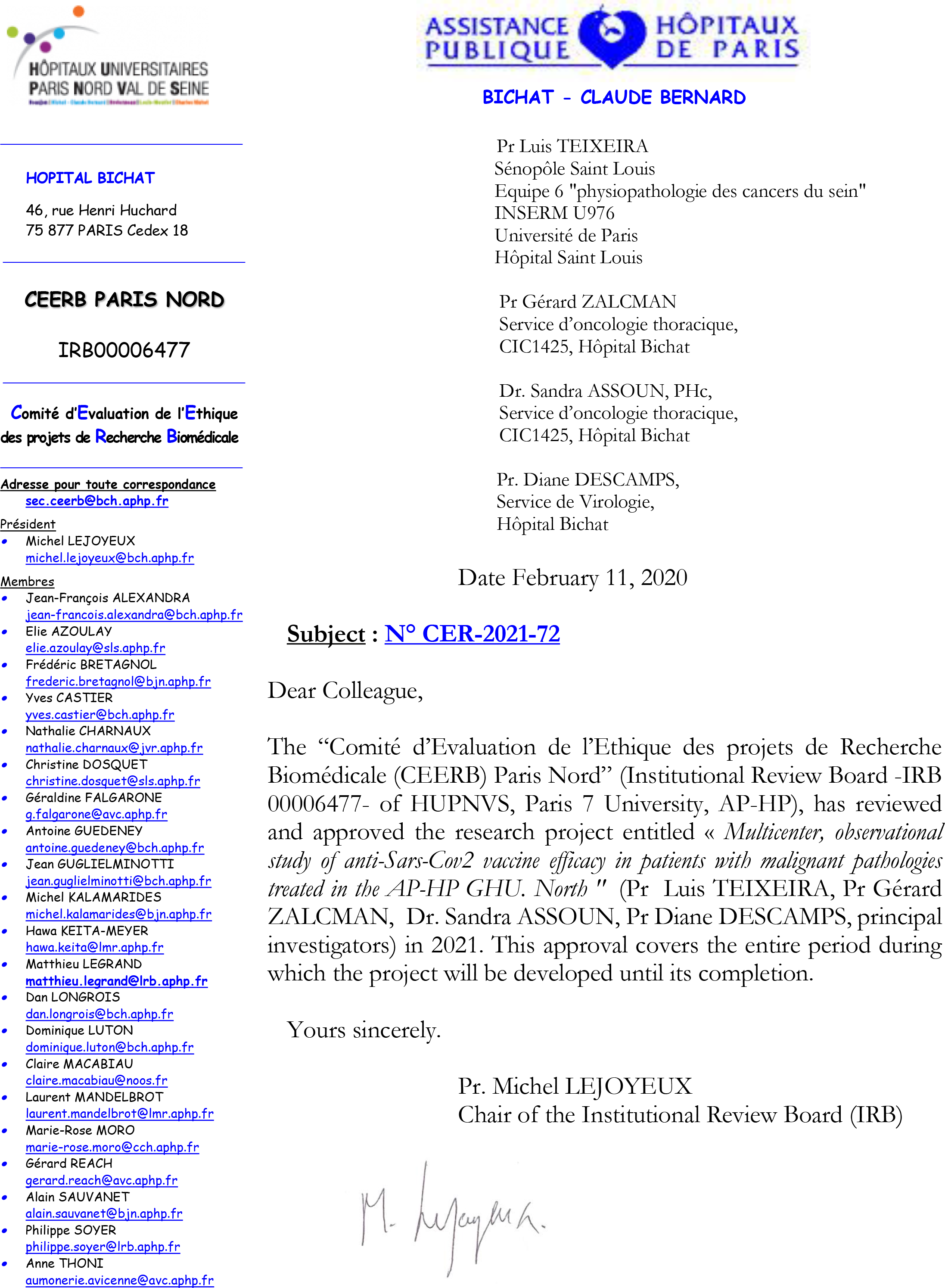

## Notes

### Competing Interest Statement

The authors have declared no competing interest.

### Clinical Trial

NCT04776005

### Funding Statement

The Assistance Publique-Hopitaux de Paris funded the study, without participating to study design, data collection, data analysis, data interpretation, or report writing.

### Author Declarations

Comite d'Evaluation de l'Ethique des projets de Recherche Biomedicale (CEERB, IRB00006477) of University Hospitals Paris-North Val-de-Seine, Paris 7 University, Assistance Publique-Hopitaux de Paris (AP-HP) upon the approval number CER-2021-72

